# Long COVID In Adults at 12 Months After Mild-to-Moderate SARS-CoV-2 Infection

**DOI:** 10.1101/2021.04.12.21255343

**Authors:** Paolo Boscolo-Rizzo, Francesco Guida, Jerry Polesel, Alberto Vito Marcuzzo, Vincenzo Capriotti, Andrea D’Alessandro, Enrico Zanelli, Riccardo Marzolino, Chiara Lazzarin, Paolo Antonucci, Erica Sacchet, Margherita Tofanelli, Daniele Borsetto, Nicoletta Gardenal, Martino Pengo, Giancarlo Tirelli

## Abstract

**Background:** In a proportion of patients recovered from the acute COVID-19 phase, a variable range of symptoms has been observed to persist for at least 6-months.

**Objectives:** The main aim of this study was to evaluate the prevalence of COVID-related symptoms 12-months after the onset of mild-to-moderate disease.

**Methods:** Prospective study based on structured questionnaires and additional outcomes.

**Results:** 304/354 patients completing the survey at baseline also completed the follow-up interview (85.9%; median [range] age, 47 [18-76] years; 185 [60.9%] women). Persistence of at least one symptom at 12-months follow-up was reported by 161 patients (53.0%). The most commonly reported symptom of long COVID was felt tired (n=83, 27.3%), followed by smell or taste impairment (n=67, 22.0%), shortness of breath (n=39, 12.8%) and muscle pain (n=28, 9.2%). Being females (OR=1.64; 95% CI: 1.00-2.70), aged between 40-54 (OR=1.92; 95% CI: 1.07-3.44), having a BMI ≥25 (OR=1.67; 95% CI: 1.00-2.78), and experiencing more symptoms during the acute phase of the disease (OR=8.71 for ≥8 symptoms; 95% CI: 2.73-27.76) were associated with long COVID. Persistence of symptoms showed a significant impact on quality of life (p<0.0001) and depression scale scores (p=0.0102).

**Conclusion:** More than half of patients with previous mild-to-moderate symptomatic COVID-19 complained the persistence of at least one symptom 12-months after the onset of the illness.

## Introduction

Since the first cases in China in December 2019, coronavirus disease 2019 (COVID-19), caused by severe acute respiratory syndrome coronavirus 2 (SARS-CoV-2) has spread globally causing a pandemic [1]. SARS-CoV-2 infection can cause a wide array of symptoms ranging from mild to severe or fatal forms with a substantial fraction of cases remaining asymptomatic carries [2]. In symptomatic subjects, the most prevalent symptoms are cough, fever, fatigue, smell or taste impairment, myalgias, and gastrointestinal symptoms [3].

Furthermore, it has been observed that in a proportion of patients recovered from the acute phase, a variable range of symptoms may persist for a long time [4,5] and the persistence of symptoms has a significant impact on health-related quality of life (QoL) [6].

There is still no consensus on how to call this condition: the names “chronic COVID”, “chronic COVID syndrome”, “long haulers”, “long COVID” have been used with the latter being the most widespread among patients and in the literature [7,8]. Also, there is a lack of a clear definition and criteria to identify this condition. Mahase defined it as: “illness in people who have either recovered from COVID-19 but are still report lasting effects of the infection or have had the usual symptoms for far longer than would be expected” [9]. A similar condition has developed during the other coronaviruses’ outbreaks, i.e., severe acute respiratory syndrome coronavirus (SARS-CoV) and Middle East respiratory syndrome coronavirus (MERS-CoV) [10].

An increasing number of studies have been focused on long COVID, but they have mainly been concentrated on previously hospitalized severe COVID-19 patients reporting symptoms up to 6-months after illness [5,11–14]. The main aim of this study was to evaluate the prevalence of COVID-related symptoms 12-months after the onset of mild-to-moderate disease.

## Materials and Methods

### Subjects

We conducted a prospective study on mild-to-moderate symptomatic adult patients consecutively assessed at Cattinara University Hospital, Trieste, Italy between March 1 and March 31, 2020, who tested positive for SARS-CoV-2 RNA by polymerase chain reaction (PCR) on nasopharyngeal and throat swabs performed according to World Health Organization recommendation [15]. All patients were initially home-isolated with mild-to-moderate symptoms. Patients were considered mildly symptomatic if they had less severe clinical symptoms with no evidence of pneumonia, not requiring hospitalization, and therefore considered suitable for being treated at home. As this study was within a research project aimed to assess the prevalence of chemosensory dysfunction in patients with SARS-CoV-2 infection[16], patients with a history of previous trauma, surgery or radiotherapy in the oral and nasal cavities, allergic rhinitis or rhinosinusitis, previous olfactory or gustative dysfunction, or psychiatric or neurological diseases, were excluded from the study.

354 (71.4%) of the 496 eligible patients completed the baseline telephone interview administrated within 3 weeks after the first positive swab performed between 1^st^ – 31^st^ March 2020. The median time from symptoms onset to SARS-CoV-2 testing was 7 days (interquartile range, 4-12 days). All patients completing the baseline interview were phoned from 5^th^ to 22^nd^ March 2021, so that all patients were recontacted 12-months after the onset of symptoms; in case of a non-response, patients were re-contacted twice.

### Questionnaires

All patients were asked the same questions. Baseline demographic and clinical data, obtained at the time of acute illness or retrospectively within 30-day from the first positive swab, were collected through *ad hoc* questions administered by telephone interview and included gender, age, self-reported height and weight, smoking and alcohol habits, and the following co-morbidities: immunosuppression, diabetes, cardiovascular diseases, active cancer, chronic respiratory disease, kidney disease, liver disease. Obesity was defined as having a body mass index (BMI) ≥ 30. Symptoms were assessed through *ad hoc* questions and structured questionnaire, i.e., the Acute Respiratory Tract Infection Questionnaire (ARTIQ). In addition, during the follow-up interview patients were asked about their general health perception and the presence of depression. Briefly, a single item was used to assess general health perceptions. Patients were asked to rate their overall health on a scale of 0-10, with 0 representing ‘worst health’ and 10 representing perfect health according to Cleary et al. [17]. Patients were also asked “on a scale of 0–10, with zero being no depression and 10 being the worst depression you can imagine, how would you rate the worst depression you have now?”[18].

### Statistical analysis

Symptom prevalence was expressed as percentage of total patients, and 95% confidence interval (CI) were calculated using Clopper-Pearson method; differences in prevalence were evaluated through Fisher’s exact test. Odds ratio (OR) of symptoms’ persistence and corresponding 95% confidence intervals (CI) were calculated through unconditional logistic regression model, adjusting for sex, age and number of symptoms during the acute Covid-19 phase. To account for potential bias of sex and age, differences in mean value of quality of life and depression were tested through the analysis of variance (ANOVA). Analyses were performed using R 3.6. and statistical significance was claimed for p<0.05 (two-tailed).

## Results

Of 354 patients completing the survey at baseline, 304 also completed the 12-months follow-up survey (85.9%). Baseline socio-demographic and clinical characteristics of 304 patients are reported in Table 1. The median age of the study cohort was 47 years (range 18-76 years). There was a female preponderance with 185 out of 304 being females (60.9%). Associated co-morbidities were reported by 98 cases (32.3%) with the most common being obesity (n=36, 11.8%) followed by cardiovascular disease (n=26, 8.6%) and chronic respiratory disease (n=21, 6.9%).

**Table 1.** Sociodemographic and clinical characteristics of 304 patients positive for COVID-19

|  | n | (%) |
| --- | --- | --- |
| Gender |  |  |
| Male | 119 | (39.1) |
| Female | 185 | (60.9) |
| Age (years)<sup>a</sup> |  |  |
| <40 | 93 | (30.6) |
| 40-54 | 119 | (39.1) |
| ≥55 | 92 | (30.3) |
| Smoking habits |  |  |
| Never | 179 | (58.9) |
| Former | 74 | (24.3) |
| Current | 51 | (16.8) |
| Drinking habits |  |  |
| Never | 202 | (66.5) |
| Former | 30 | (9.9) |
| Current | 72 | (23.7) |
| BMI (kg m<sup>-2</sup>) |  |  |
| <25 | 170 | (55.9) |
| ≥25 | 134 | (44.1) |
| Comorbidity |  |  |
| None | 206 | (67.8) |
| Any | 98 | (32.2) |
| Obesity | 36 | (11.8) |
| Cardiovascular disease | 26 | (8.6) |
| Chronic respiratory disease | 21 | (6.9) |
| Immunosuppression | 18 | (5.9) |
| Diabetes | 10 | (3.3) |
| Cancer | 5 | (1.6) |
| Liver disease | 5 | (1.6) |
| Kidney disease | 2 | (0.7) |
| Time for a negative swab (days) |  |  |
| ≤23 | 157 | (51.6) |
| >23 | 147 | (48.4) |

BMI: *body mass index*<sup>a</sup>Median age (IQR): 47 years (37-56)

The median time to achieving a negative swab was 23 days (interquartile range, 15-32 days). Figure 1 and Supplementary Table 1 report the prevalence of symptoms in the acute phase of COVID-19 and at 12-months follow-up. During the acute phase of COVID-19 the most frequently reported symptoms were fever (n=232, 76.3%), felt tired (n=216, 71.1%), smell impairment (n=201, 66.1%), taste impairment (n=198, 65.1%) and joint pain (n=181, 59.5%).

**Figure 1.**
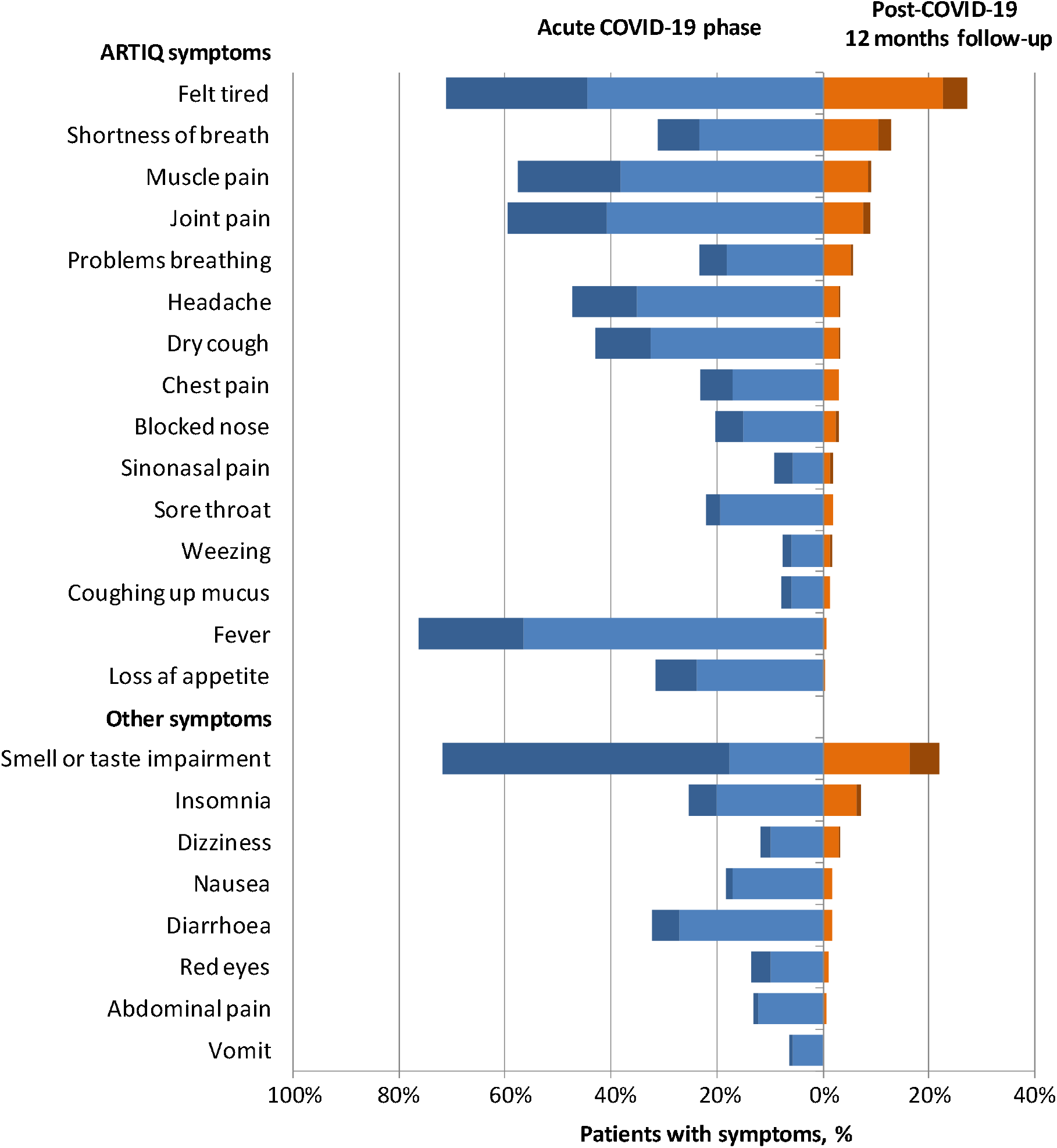
COVID-19-Related Symptoms. The figure reports the percentages of patients with COVID-19-related symptoms during the acute phase of illness (blue) and at 12-months follow-up (orange) according to severity (light=mild; dark=severe) ARTIQ: *Acute Respiratory Tract Infection Questionnaire*

Persistence of at least one symptom at 12-months follow-up was reported by 161 patients (53.0%). The symptom that most frequently persisted at 12-months was felt tired (n=83, 27.3%), followed by smell or taste impairment (n=67, 22.0%) with smell being altered in 62 patients (20.4%) and taste in 46 (15.1%), shortness of breath (n=39, 12.8%) and muscle pain (n=28, 9.2%).

The risk of symptom’s persistence at 12-months follow-up was trendy higher in females (OR=1.64; 95% CI: 1.00-2.70; p=0.0505), and significantly higher in subjects aging between 40-54 (OR=1.92; 95% CI: 1.07-3.44; p=0.0290) and in those with a BMI ≥25 (OR=1.67; 95% CI: 1.00-2.78; p=0.0492). The presence of 3 to 7 symptoms during the acute phase of the disease resulted associated with a significant higher odd of symptom’s persistence after 12-months (OR=3.22; 95% CI: 1.01-10.24; p=0.0475). This association resulted stronger in those having ≥8 symptoms (OR=8.71; 95% CI: 2.73-27.76; p=0.0003) (Table 2).

**Table 2.**
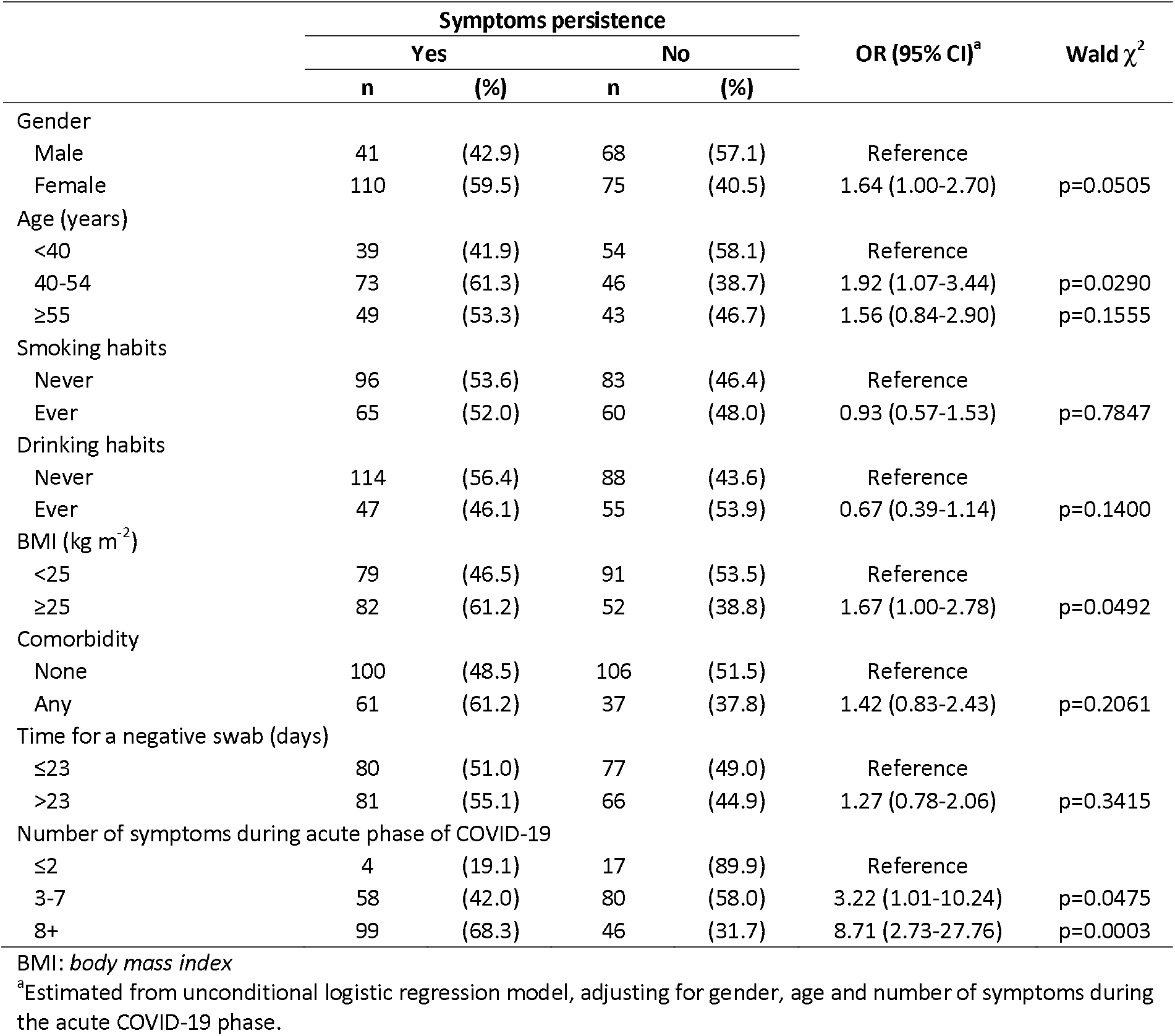
Odds ratio and corresponding 95% confidence interval for symptoms persistence at 12-months follow-up according to sociodemographic and clinical characteristics in 304 patients positive for COVID-19

In long haulers, 8 patients (5.0%; 95% CI: 2.2-9.6%) had QoL scores ≤5 and 23 patients (14.3%; 95% CI: 9.3-20.7%) had a depression score ≥5. Persistence of symptoms 12-months after the onset was associated with worse quality of life (p<0.0001) and worse scores on depression scale (p=0.0102). In particular, quality of life was significantly affected by felt tired (p<0.0001), shortness of breath (p=0.0015) and insomnia (p=0.0379), while felt tired significantly impact on scores on depression scale (p=0.0082) (Table 3).

**Table 3.** Mean value and standard deviation (std) of quality of life (0=the worst; 10=the best) and depression (0=the best; 10=the worst) at 12-months follow-up in 304 patients COVID-19

|  | Quality of life |  | Depression |  |
| --- | --- | --- | --- | --- |
|  | Mean (std) | ANOVA<sup>a</sup> | Mean (std) | ANOVA<sup>a</sup> |
| Gender |  |  |  |  |
| Male | 8.39 (1.12) | p=0.0787 | 1.98 (1.63) | p=0.0339 |
| Female | 8.04 (1.61) |  | 2.52 (2.17) |  |
| Age (years) |  |  |  |  |
| <40 | 8.65 (1.18) | p=0.0030 | 2.10 (1.68) | p=0.4489 |
| 40-54 | 8.01 (1.55) |  | 2.54 (2.06) |  |
| ≥55 | 7.92 (1.46) |  | 2.24 (2.17) |  |
| Comorbidities |  |  |  |  |
| None | 8.29 (1.58) | p=0.2125 | 2.27 (2.01) | p=0.3487 |
| Any | 7.95 (1.09) |  | 2.49 (1.93) |  |
| Persistent symptoms |  |  |  |  |
| None | 8.65 (1.30) | p<0.0001 | 1.94 (1.85) | p=0.0102 |
| Any | 7.75 (1.44) |  | 2.64 (2.06) |  |
| Persistent shortness of breath |  |  |  |  |
| No | 8.30 (1.44) | p=0.0015 | 2.25 (1.95) | p=0.3525 |
| Yes | 7.36 (1.20) |  | 2.72 (2.24) |  |
| Persistent felt tired |  |  |  |  |
| No | 8.45 (1.28) | p<0.0001 | 2.10 (1.79) | p=0.0082 |
| Yes | 7.46 (1.62) |  | 2.87 (2.37) |  |
| Persistent insomnia |  |  |  |  |
| No | 8.23 (1.46) | p=0.0379 | 2.26 (1.98) | p=0.1429 |
| Yes | 7.50 (1.10) |  | 3.00 (2.07) |  |
| Persistent impairment of sense of smell or taste |  |  |  |  |
| No | 8.26 (1.46) | p=0.0532 | 2.22 (2.05) | p=0.2084 |
| Yes | 7.88 (1.39) |  | 2.63 (1.75) |  |

<sup>a</sup>Analysis of Variance (ANOVA) was performed through generalized linear model for unbalanced data adjusted from gender and age

## Discussion

To our knowledge, this is the cohort study with longest follow-up duration assessing the prevalence of persistent symptoms in COVID-19 patients. We observed that 12-months after the onset of illness, 53.0% of patients with mild-to-moderate COVID-19 endorsed at least one persistent symptom. A previous investigation in a large cohort of patients with COVID-19 discharged from hospital reported a at least one symptom in 76% of cases 6-months after acute infection [12], while in another study, including outpatients with mild disease, approximately 30% reported persistent symptoms at 6-months [14]. Thus, taking into account that our series included only outpatients with mild-to-moderate COVID-19 representing the overwhelming majority of COVID-19 patients [3], the rate of long COVID observed in the present cohort is quite worrying. Although few patients had QoL scores ≤5 and depression score ≥5, the persistence of symptoms 12-months after the onset was associated with worse quality of life and worse scores on depression scale. Recent investigations have observed a significant impairment in QoL in patients previously hospitalized for COVID-19 [6] and a worsening of QoL was described in about one third of patients 6-months after the acute illness [14]. Considering the continued worldwide spread of SARS-CoV-2 infection, the burden of long haulers on the health care systems will be pressing and urged researchers to identify subjects at risk of long COVID for trials of strategies for prevention or treatment and to plan education and rehabilitation services in order to face with the considerable health and economic concerns [4].

In agreement with other evidence, the most commonly reported symptom was fatigue followed by chemosensory dysfunction [4,5,14]. Fatigue was reported by 27% of patients. Previous investigations with long-term follow-up have observed a frequency of this symptom of 14% [14] to 63% [12] at 6-months with higher prevalence recorded in a series including only hospitalized subjects. An altered sense of smell or taste was the second more common symptom of long COVID in the present series reported by 22.0% of subjects. Chemosensory dysfunction has been consistently reported as one of the most predominant symptom during the acute phase of the disease as well as among the most frequent long-lasting symptoms in patients with COVID-19 [19,20] with a higher prevalence in patients with mild-to-moderate disease [21]. Consistently, series including outpatients [4,14] have observed a higher rate of long-term chemosensory impairment compared to those including only patients with severe COVID-19 [12], while one study not using structured questionnaires failed to provide data about the prevalence of altered sense of smell or taste in the context of long COVID syndrome [11]. Furthermore, anosmia was recently reported as one of the symptoms characterizing the long COVID syndrome among a large series of app users [4]. Some authors have speculated that mild-to-moderate COVID-19 patients may be affected by nasal-centric viral spread, whereas patients requiring hospitalization may be experiencing a more pulmonary-centric and systemic viral infection thus impacting the prevalence of different symptoms both in acute phase and in the context of long COVID syndrome [22]. However, although all patients in the study had mild to moderate disease without evidence of pneumonia, 13% still reported shortness of breath at 12-months follow-up interview. Interestingly, an abnormal pulmonary function consisting in an impaired diffusing-capacity was observed in 30% of patients after the acute phase of mild COVID-19 without pneumonia [23].

In addition to the different settings and timing of symptoms detection, an important source of heterogeneity in reporting the prevalence of symptoms in patients with long COVID is the lack of using a validated questionnaire. To date, no standardized tool has been proposed yet for a uniform and comprehensive evaluation and reporting of symptoms in COVID-19 patients. In the present study *ad hoc* questions were used as well as a structured questionnaire, the ARTIQ, as it was demonstrated to be a validated tool to assess the severity and functional impacts of acute respiratory tract infections [24].

Experiencing more symptoms at baseline was the most significant factor associated with long COVID. This is in line with recent evidence showing that subjects complaining more than 5 symptoms during the first week of illness are more prone to develop long COVID [4]. Moreover, the prevalence of long haulers was higher in females, patients in their middle age and those with BMI ≥25. Thus, models to identify patients at risk for long COVID may be developed.

This study has several limitations. First, the absence of a control group may impact on the interpretation of results. The single-centre study design and the relatively small number of sample size may restrict its generalizability. Hospitalized patients were not included in the study. Although this made our cohort more homogeneous, studies comparing the long-term outcomes in inpatients and outpatients are definitely needed. Symptoms were self-reported and based on telephone interview. Though, we tried to perform a comprehensive symptoms assessment by using *ad hoc* questions and a structured questionnaire, some symptoms may have been undetected. The QoL and depression assessment was performed only at 12-months follow-up. Thus, we were unable to report a worsenig in the QoL and depression scale scores.

In conclusion, our study indicates that persistent symptoms of SARS-CoV-2 infection can be detected beyond 12-months from the onset of the illness in more than half of patients with mild-to-moderate disease. Identifying patients at risk for prevention and treatment will be critical to improving outcomes and reducing health costs. Finally, a structured and validated questionnaire for the assessment of symptoms in COVID-19 patients is highly desirable to characterize the full clinical spectrum of long COVID and improve the reliability and reproducibility of clinical studies.

## Data Availability

The authors confirm that the data supporting the findings of this study are available
within the article

## Funding

None

## Conflict of interest

The authors declare that they have no conflicts of interest

## Availability of data and material

The authors confirm that the data supporting the findings of this study are available within the article

## Code availability

not applicable

## Author Contributions

Drs Boscolo-Rizzo and Guida had full access to all of the data in the study and take responsibility for the integrity of the data and the accuracy of the data analysis

*Concept and design*Drs Boscolo-Rizzo, Guida, Pengo and Tirelli

*Acquisition, analysis, or interpretation of data*: Drs Boscolo-Rizzo, Guida, Polesel, Marcuzzo, Antonucci, Capriotti, Sacchet, D’Alessandro, Zanelli, Marzolino, Lazzarin, Tofanelli, Gardenal, Borsetto

*Drafting of the manuscript*: Drs Boscolo-Rizzo, Guida, Polesel, Pengo, Tirelli

*Critical revision of the manuscript for important intellectual content*: Drs Boscolo-Rizzo, Guida, Polesel, Borsetto, Pengo, Tirelli

*Statistical analysis*: ⍰Polesel

*Supervision:* Drs Boscolo-Rizzo and Tirelli

## Ethics approval

The study was approved by the Ethics Committee of the Friuli Venezia Giulia Region (CEUR-2020-Os-156)

## Informed consent

Additional informed consent was obtained from all individual participants for whom identifying information is included in this article

---

**Supplementary Table 1.** Evolution of COVID-19-related symptoms in 304 patients

| Symptoms | Post-COVID-19 12-months follow-up |  | Acute COVID-19 phase |  |
| --- | --- | --- | --- | --- |
|  | n | % (95% CI) | n | % (95% CI) |
| **Any symptom** | 161 | 53.0 (47.2-58.7) | 304 | 100 (98.8-100) |
| **ARTIQ** |  |  |  |  |
| Felt tired | 83 | 27.3 (22.4-32.7) | 216 | 71.1 (65.6-76.1) |
| Shortness of breath | 39 | 12.8 (9.3-17.1) | 95 | 31.3 (26.1-36.8) |
| Muscle pain | 28 | 9.2 (6.2-13.0) | 175 | 57.6 (51.8-63.2) |
| Joint pain | 27 | 8.9 (5.9-12.7) | 181 | 59.5 (53.8-65.1) |
| Problems breathing | 17 | 5.6 (3.3-8.8) | 71 | 23.4 (18.7-28.5) |
| Dry cough | 10 | 3.2 (1.6-6.0) | 131 | 43.1 (37.5-48.9) |
| Headache | 10 | 3.2 (1.6-6.0) | 144 | 47.4 (41.6-53.1) |
| Blocked nose | 9 | 3.0 (1.4-5.5) | 62 | 20.4 (16.0-25.4) |
| Chest pain | 9 | 3.0 (1.4-5.5) | 70 | 23.0 (18.4-28.2) |
| Sore throat | 6 | 2.0 (0.7-4.2) | 67 | 22.0 (17.5-27.1) |
| Sinonasal pain | 6 | 2.0 (0.7-4.2) | 28 | 9.2 (6.2-13.0) |
| Weezing | 5 | 1.6 (0.5-3.8) | 23 | 7.6 (4.9-11.1) |
| Coughing up mucus | 4 | 1.3 (0.4-3.3) | 24 | 7.9 (5.1-11.5) |
| Fever | 2 | 0.7 (0.1-2.4) | 232 | 76.3 (71.1-81.0) |
| Loss of appetite | 1 | 0.3 (0.0-1.8) | 96 | 31.6 (26.4-37.1) |
| **Other symptoms** |  |  |  |  |
| Smell impairment | 62 | 20.4 (16.0-25.4) | 201 | 66.1 (60.5-71.4) |
| Taste impairment | 46 | 15.1 (11.3-19.7) | 198 | 65.1 (59.5-70.5) |
| Insomnia | 22 | 7.2 (4.6-10.8) | 77 | 25.3 (20.5-30.6) |
| Dizziness | 10 | 3.2 (1.6-6.0) | 36 | 11.8 (8.4-16.0) |
| Diarrhoea | 5 | 1.6 (0.5-3.8) | 98 | 32.2 (27.0-37.8) |
| Nausea | 5 | 1.6 (0.5-3.8) | 56 | 18.4 (14.2-23.2) |
| Red eyes | 3 | 1.0 (0.2-2.9) | 41 | 13.5 (9.9-17.8) |
| Abdominal pain | 2 | 0.7 (0.1-2.4) | 40 | 13.2 (9.6-17.5) |
| Vomit | 0 | 0.0 (0.0-1.2) | 19 | 6.3 (3.8-9.6) |

ARTIQ: *Acute Respiratory Tract Infection Questionnaire*

## Notes

### Competing Interest Statement

The authors have declared no competing interest.

### Author Declarations

The study was approved by the Ethics Committee of the Friuli Venezia Giulia Region (CEUR-2020-Os- 156)

